# Rt.GLM: Unifying estimation of the time-varying reproduction number, *R_t_*, under the Generalised Linear and Additive Models

**DOI:** 10.1101/2025.06.24.25330176

**Authors:** Pierre Nouvellet

## Abstract

Most current methods to estimate the time-varying reproduction number (R_t_), such as *EpiEstim*, rely on branching processes and the renewal equation. They also require subjective choices to set the level of temporal and spatial heterogeneity assumed. We propose a novel framework to estimate R_t_ based on Generalized Linear and Additive Models (GLM/GAM). By integrating the renewal equation model within GLM/GAM, the proposed framework, “*Rt.glm*”, allows smooth estimation of R_t_ variations over time and space without relying on arbitrary scaling parameters.

The performance of *Rt.glm* was evaluated using historical datasets and simulated outbreaks. It demonstrated improved overall performance and accuracy compared to *EpiEstim*, as measured by the CRPS scores and Mean Square Errors respectively. However, when case incidence was low and *R_t_* estimation relied on a smoothing term, *Rt.glm* was marginally overconfident in its estimates.

The method offers substantial improvement for the real-time estimation of spatio-temporal trends in *R_t_*, with improved performance and lower reliance on arbitrarily set parameters. The open-source and user-friendly R package developed will also simplify user experience. Finally, the framework bridges gaps between epidemic monitoring methodologies and sets the stage for future extensions to enhance statistical inference and integrate additional epidemiological complexities, including the evaluation of intervention strategies.

**Highlights:**

- A novel framework is introduced to estimate R_t_ using GLM and GAM approaches.
- This allows smooth spatio-temporal estimation of R_t_ without predefined scales.
- Overall, it outperforms *EpiEstim*, with lower Mean Square Error and CRPS scores.
- An open-source, user-friendly R package is provided for real-time R_t_ estimation.
- This proof-of-concept provides a strong foundation for future developments.

## Introduction

The past decade has seen a surge of interest in infectious disease epidemiology research, particularly in the context of emerging pathogens(1–6). Much of this interest focuses on estimating transmissibility, often measured through the reproduction number: the average number of secondary infections generated by an infected individual(7). While crucial to monitor the spread and intensity of transmission (8,9), accurate estimation of transmissibility can also improve our understanding of the impact of interventions on transmission(10,11), opening the way for an evidence-based response(12,13). Thus, the concept of the time-varying reproduction number, *R_t_* - or “the R number” as often referred to in the media during the COVID-19 pandemic - has become a widely popular concept.

Estimating *R_t_* is not a new challenge, and in the early 2000s, approaches to estimate *R_t_* from epidemiological data were developed (14,15), typically relying on the number of new cases reported over time - as serological, household surveys or other sources of information are rarely available during an emerging epidemic. Methods quickly built on branching process theory within the ‘renewal equation’ framework (7,16,17), which underpinned a wealth of statistical methods. From the 2009 pandemic influenza’s emergence (18) to numerous Ebola epidemics in West (1) and Central Africa(19), as well as the COVID-19 pandemic(20), these methods have proven critical to monitor epidemics, evaluate the impact of interventions and communicate the risk of transmission to policymakers and the public.

However, as *R_t_* can fluctuate over time and space, estimating and monitoring this parameter is challenging. Although widely used, the current ‘renewal equation’ framework and associated methods still do not fully address two important questions regarding the *temporal and spatial scale of heterogeneities* in *R_t_*, i.e. how swiftly transmission changes over time and /or space. For instance, when using the *EpiEstim* R-package(16,21), arguably one of the most popular and performant existing tool, users must define the level of temporal and spatial granularity by assuming 1) a constant *R_t_* during pre-specified time-windows, 2) aggregating case counts spatially and assuming *R_t_* are independent across locations. While *EpiEstim*’s computational efficiency makes it possible to run extensive sensitivity analyses, assuming various levels of spatial and temporal aggregation, comparing the resulting estimates and fits is not trivial. In practice, the choice of temporal and spatial scales is therefore typically guided more by experience and data availability rather than a sound statistical rationale.

The limitations mentioned above are well described in the literature, and some of them have been addressed. For instance, *Epidemia* was instrumental in pulling information from multiple locations to infer drivers of transmissibility(20), while *EpiLPS* made a key contribution in allowing the smoothing (B-spline) of temporal variation in *R_t_* (22). However, no framework currently tackles both challenges in a validated, intuitive and user-friendly tool.

Here, we first propose a novel framework to estimate *R_t_* that borrows from the renewal equation framework and the ‘classical’ GLM/GAM statistical framework (Generalised Linear/Additive Models). We then evaluate its performance on real and simulated data to characterise temporal and spatial heterogeneities in transmission, using a newly developed R-package (*Rt.glm*). This contribution is intended to be a proof-of-concept, demonstrating 1) the equivalence of renewal-based methods and GLMs to estimate transmissibility of a pathogen, and 2) some of the key possibilities offered by this novel framework.

## Methods

### a) General approach

The general approach relies on a simple observation: most *R_t_* estimation tools (10,11,20) rely on the ‘renewal equation’ framework(7), and assume that the incidence at time ‘t’, *I*_*t*_, is Poisson distributed:

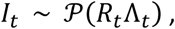

Where 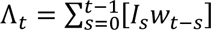 is the sum of past incidence weighted by the infectivity function *w*. In practice, the infectivity function is taken to be the Generation Time (GT) distribution: the time between infection of infector-infectee pairs. As infection times are usually unobserved, The serial interval (the time between *onset of symptoms*) is commonly used in practice as a proxy for the GT.

First, note that Λ_*t*_ is parameter-free: it can be pre-computed prior to inference. Therefore, the estimation of *R_t_* above can be framed as a GLM (Generalized Linear Model), with Λ_*t*_ viewed as an offset within the GLM framework:

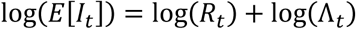

#### Estimating Rt using Generalised Linear Model (GLM)

Thus, regressing *I_t_* against time, without intercept, with log(Λ_*t*_) as an offset, within a GLM/GAM assuming a Poisson distribution with log link function, should result in an inference equivalent to that implemented in the *EpiEstim* R-package.

This results in a simple R-code to implement a GLM version of EpiEstim (where Λ_*t*_ is precomputed as described above):

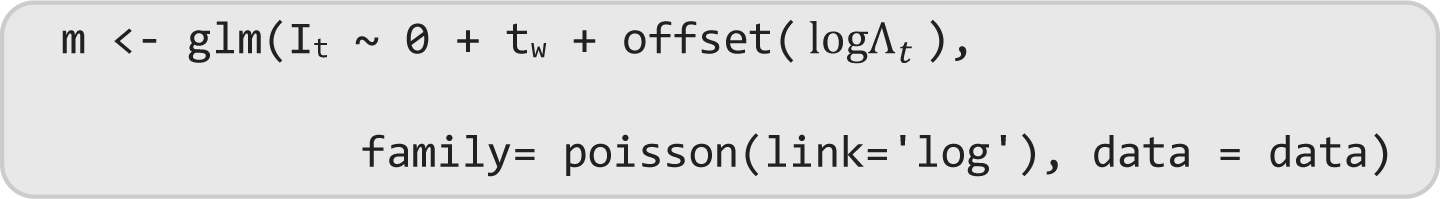

where **data** above is an R dataframe containing all relevant variables as columns (i.e. `I, t`_w_ `and` logΛ_*t*_). In the above code, ‘`t`_w_ ‘ is a vector of indexes (a fixed factor in the GLM language) specifying which time window each time step belongs. As in *EpiEstim*, during each time window, *R_t_* is assumed constant. Note that the equivalent concept of time-window within the *EpiEstim* framework, is implemented slightly differently with user setting the start and end dates of each time-window. The coefficients returned in ‘`m`’ give us point estimates of *R_t_*, and 95% Confidence Intervals (95%CI), both of which may be compared to the central estimates and 95%Credible Intervals returned by *EpiEstim*.

In *EpiEstim*, the time-windows can be non-overlapping, i.e. one *R_t_* is estimated per time-window. Alternatively, time-windows may be overlapping, a process akin to estimating a moving average of *R_t_*, i.e. one *R_t_* is estimated per time-window but observed incidence data may contribute to multiple time-windows and therefore contribute to multiple *R_t_* ‘s. The process of estimating *R_t_* with overlapping time-windows can be reproduced using our GLM framework by repeating incidence observations in the ‘data’ dataframe and assigning the time windows appropriately.

The approach described not only potentially simplifies the inference compared to EpiEstim, from a user-perspective, but more importantly, it opens avenues for extending the framework, building on decades of work on GLMs. Specifically, here we will demonstrate two extensions relying on the development of General Additive Models (GAMs)(23) to tackle challenges linked to uncertainties in temporal and spatial variations of *R_t_*.

In situations where more than one location, ‘l’, is considered, the framework above can hold:

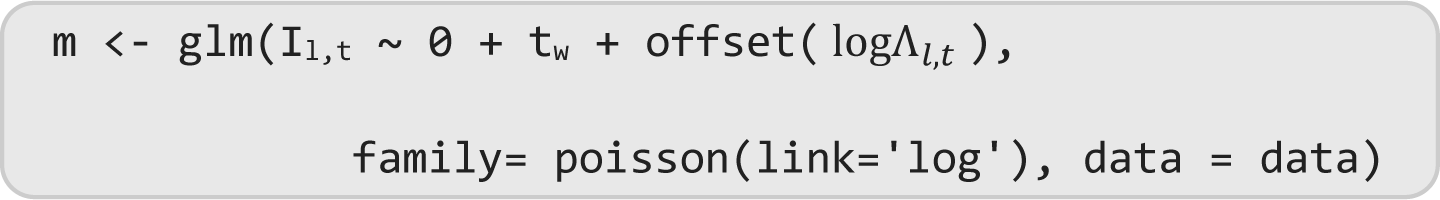

where `I`_l,t_ `, t`_w_ `a`nd logΛ_*l*,*t*_ are vectors specifying the incidences observed, time-windows indexes and offsets for each location and time step.

Note that currently, in the EpiEstim framework, estimations of *R_t_* ‘s for each location need to be performed independently, while in our framework, all estimations can be performed jointly.

#### Estimating Rt using Generalised Additive Model (GAM)

A GAM is an extension of GLMs where the outcome (or more precisely the outcome transformed by the link function) may depend linearly on unknown smooth functions of some predictor variables.

Therefore, by including a smooth temporal term, one can infer temporal variations in *R_t_* without specifying the scale of temporal smoothing, as this is internally inferred within a sound statistical framework(24) (see Supplementary Information section 1-SI1- for details on the parameterisation of the GAM). A simple implementation using the *mgcv* R-package(23) would be:

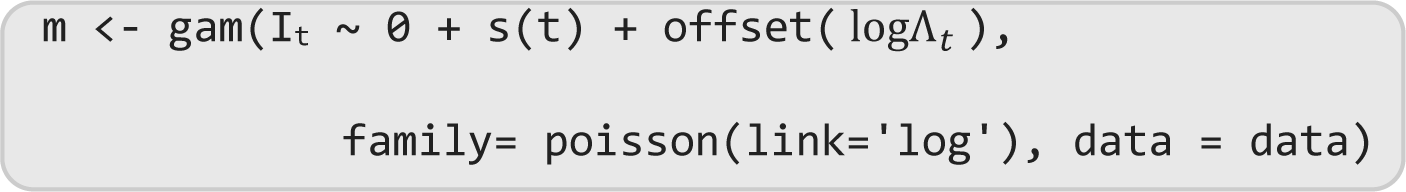

Additionally, when multiple locations are considered, by including a smooth spatial term (e.g. latitude, longitude - lat/long - of spatial units), one can infer spatial variations in *R_t_* without specifying the spatial scale at which incidence needs to be aggregated (again see SI1 for details and discussion on parameterisation):

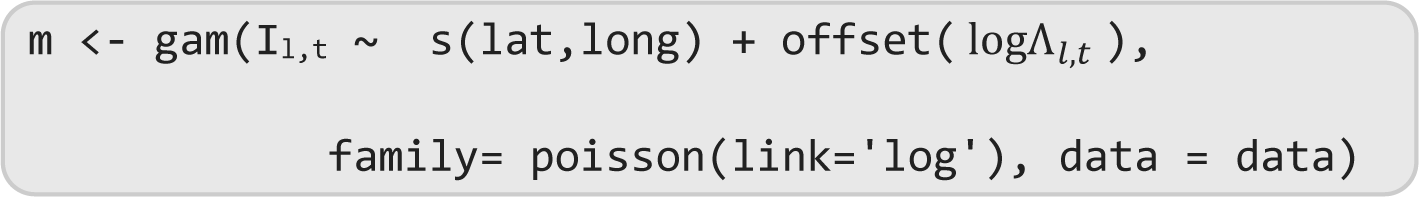

An R package, *Rt.glm* to implement the Rt-GLM and Rt-GAM frameworks, facilitates estimations including computation of Λ_*t*_, implementation of overlapping time windows and provides *R_t_* estimates with 95%CI as output. The package is available on github (https://github.com/pnouvellet/Rtglm).

### b) Comparing estimates of Rt

#### i. Estimating R_t_ with GLM

To validate our novel framework, we compare *R_t_* estimates using *EpiEstim* vs. our *Rt.glm* framework.

The first comparison relies on datasets available within the *EpiEstim* R-package, i.e. datasets including daily reported incidence data (*I*_*t*_) and an associated serial interval distribution (*w*_*t*_) for: a) a 1861 measles epidemic in Hagelloch, Germany, b) a 1918 H1N1 influenza epidemic in Baltimore, c) a 1972 smallpox epidemic in Kosovo, d) the 2003 SARS epidemic in Hong Kong, and e) a 2009 H1N1 influenza outbreak in a school in Pennsylvania.

For *EpiEstim*, we chose a weakly informative prior for *R_t_* (i.e. the default mean and standard deviation of *R_t_* priors of 5). We compare results when estimating *R_t_* with overlapping, as well as non-overlapping, time-windows of size 3, 7 and 15 days.

In the main text, we present results of *R_t_* estimations using *EpiEstim* vs. *Rt.glm* for the 1918 H1N1 influenza and the 1972 smallpox epidemic in Kosovo datasets associated with time-windows of 7 days. Other scenarios for the same datasets (e.g. distinct time-windows), as well as similar comparisons using the three alternative datasets are presented in the SI. Finally, also in SI, and using the 1918 H1N1 influenza data, we briefly present the ability of *Rt.glm* to estimate *R_t_* with a temporal smooth term.

After demonstrating the equivalence of estimating *R_t_* using the *EpiEstim* or *Rt.glm* frameworks, we extend our application of *Rt.glm*, relying on GAM, to recover temporal and spatial trends in *R_t_*. We benchmarked the performance of both frameworks relying on simulated dataset.

#### ii. Estimating temporal variation in Rt using GAM

Assuming a sinusoidal *R_t_* with an amplitude ranging from 0.8 and 1.2, and a period of 30 days, we simulated 1,000 stochastic incidence curves for 100 days. The simulated incidence curves were generated using the renewal equation (as above), assuming the serial interval distribution for influenza (25), which is associated with the 1918 H1N1 influenza epidemic in Baltimore in the *EpiEstim* R-package and 30 initial cases on day 0. Simulations were obtained using the *projections* R-package(16).

Using *EpiEstim*, we then estimated *R_t_* over time using overlapping, as well as non-overlapping, time-windows of 3 and 7 days. Similar estimations of *R_t_* were obtained using our *Rt.glm* framework and package, using a GAM estimation that included a smooth temporal term (i.e. thin plate spline, see SI1 for details on GAM parameterisation). The GAM estimates relied on the *mgcv* R-package(23).

For each simulated incidence curve, we obtained 5 *R_t_* estimates over time: 4 estimates of *R_t_* temporal trends based on *EpiEstim* (i.e. 2 time-windows x overlapping/non-overlapping time-windows), and 1 estimate of *R_t_* temporal trend based on *Rt.glm*. For each, we evaluated the performance based on the Mean Square Error between the estimated *R_t_* and that used for simulation (MSE), the proportion of estimated 95% CI (or CrI) that included the true values of *R_t_* used for simulations (coverage) and the mean Continuous Ranked Probability Score (CRPS). The CRPS metric is s a ‘strictly proper scoring rule’(26), much used in meteorology, and increasingly used in infectious disease epidemiology. CRPS measures the distance between the distribution of the true values and that of the estimated ones (27). Lower CRPS score indicates a better fit, and the score penalises both over- and under-confidence. The score is also stable (i.e. not sensitive) in handling outliers. CRPS was estimated using the *scoringutils* R-package.

We present results across simulations in summary tables, and visually checked the performance for various incidence simulated. For presentation, we visualise performance for 4 selected simulations (one figure in the main and the 3 remaining in the SI). To choose the simulations visually presented, we sorted the simulations by decreasing quality of the fit (measured by CRPS) and chose 4 that led to the 25% best, the median best, the 75% best and the worst *Rt.glm* fit. In the main text, we present the results for the simulated incidence that led to the 25% best *Rt.glm* fit, while in the SI, we present fit based on the simulated incidence that led to the median and 75% best *Rt.glm* fit, as well as the worst *Rt.glm* fit.

This strategy was chosen as the link between a stochastically simulated incidence trajectory and the resulting *R_t_* estimates is essential to assess the performance of the methods, i.e. presenting all incidence simulated and all the *R_t_* estimates would effectively loose the information on which incidence links to which *R_t_* estimates.

#### iii. Estimating Spatial variation in Rt using GAM

First, we simulated 100 independent kernels of *R_t_* across the 183 Unitary Authorities (UA) of the United Kingdom (UK) (see(28) for UA boundaries). Each kernel was produced by the superposition of 3 bi-variate Normal distributions. For each distribution, the mean (i.e. peak) was randomly assigned to the centroid of one UA, while the variance was randomly chosen from a uniform distribution ranging from 0.5 to 2 (resulting in narrow, or wide peak respectively). Each kernel was then normalised (to add to 1), and rescaled to ensure that across the UK, *R_t_* varied between 0.5 and 2.

For each location, incidence over a period of 5 days was then simulated, and when estimating *R_t_*, transmission level was assumed constant over this time period (e.g. a single time-window per location). Therefore, no temporal variation was included in either the simulation or the estimation process.

For each of the 100 *R_t_* kernels, we simulated 100 stochastic incidence curves. Again, the simulated incidence curves were generated using the *projections* R-package, assuming the serial interval distribution of the 1918 H1N1 influenza epidemic in Baltimore available from the *EpiEstim* R-package. Simulating a short incidence time-series (i.e. 5 days) with a relatively long serial interval can lead to discontinuities in incidence. To avoid this, we back-calculated the expected growth rate (29) for each UA, given the local simulated *R_t_* and assumed serial interval distribution. We then used this growth rate to compute the expected incidence for the previous 10 days (i.e. prior to the inference period), such that if *R_t_* was 1 across all UA, we would expect 30 cases in each location on day 1 of the simulation. This strategy was used to simulate incidence data on similar scale as that simulated in section (ii) above. While inference relied on those pre-inference cases (i.e. to compute Λ_*t*_), only the forward 5-days incidence simulation was used as an outcome to estimate *R_t_*.

As in the previous section, *R_t_* across UA was estimated using *EpiEstim* and *Rt.glm*.

For EpiEstim estimates, first, each UA incidence data was used independently (EpiEstim Rt – UA). As an alternative, one could assume some level of spatial homogeneity where *R_t_* is assumed constant. To mimic this, the incidences of UAs belonging to the same homogenous units were aggregated to obtain a single incidence time series and a single *R_t_* estimate. We divided the UK into a grid of 14 units. Each UA was assigned to this grid based on its centroid. Those estimates of *R_t_*, which assume homogeneity within a grid, are referred to as ‘EpiEstim Rt – medium grid’.

Finally, we obtained spatial *R_t_* estimates using a GAM estimation and including a smooth spatial term (i.e. thin plate spline, see SI1 for details on GAM parameterisation), relying on the *mgcv* R-package (23).

As previously, each spatial *R_t_* estimates were evaluated based on MSE, coverage and CRPS.

Again, we present performance across simulations in summary tables, and visually checked performance for various incidences simulated. As for temporal trends, in the main text, we present the results for the simulated incidence that led to the 25% best *Rt.glm* fit (based on the ranked CRPS scores). In the SI, we present fit based on the simulated incidence that led to the median and 75% best *Rt.glm* fit, as well as the worst *Rt.glm* fit.

## Results

### a) Estimating Rt with GLM

We confirmed that we could largely reproduce estimates from *EpiEstim* using our *Rt.glm* framework for multiple pathogens and situations. Figure 1 presents Rt estimates for the 1918 influenza in Baltimore (a-c) and the 1972 smallpox epidemic in Kosovo (d-f) using a 7-day time-window with overlapping (b,e) and non-overlapping time-windows (c,f). See SI for alternative datasets and time-window lengths. We found excellent agreement between *EpiEstim* (black) and *Rt.glm* (red), with central estimates (and 95%CI) almost exactly overlapping and that were virtually indistinguishable from each other in most situations. However, estimates could differ, especially when incidence or rather the overall infectivity (Λ_*t*_) was low (Fig. 1.e-f for instance). When this happened, the greatest discrepancy involved large confidence intervals associated with *Rt.glm*, while the confidence intervals returned by *EpiEstim* reflected the prior distribution.

**Figure 1:**
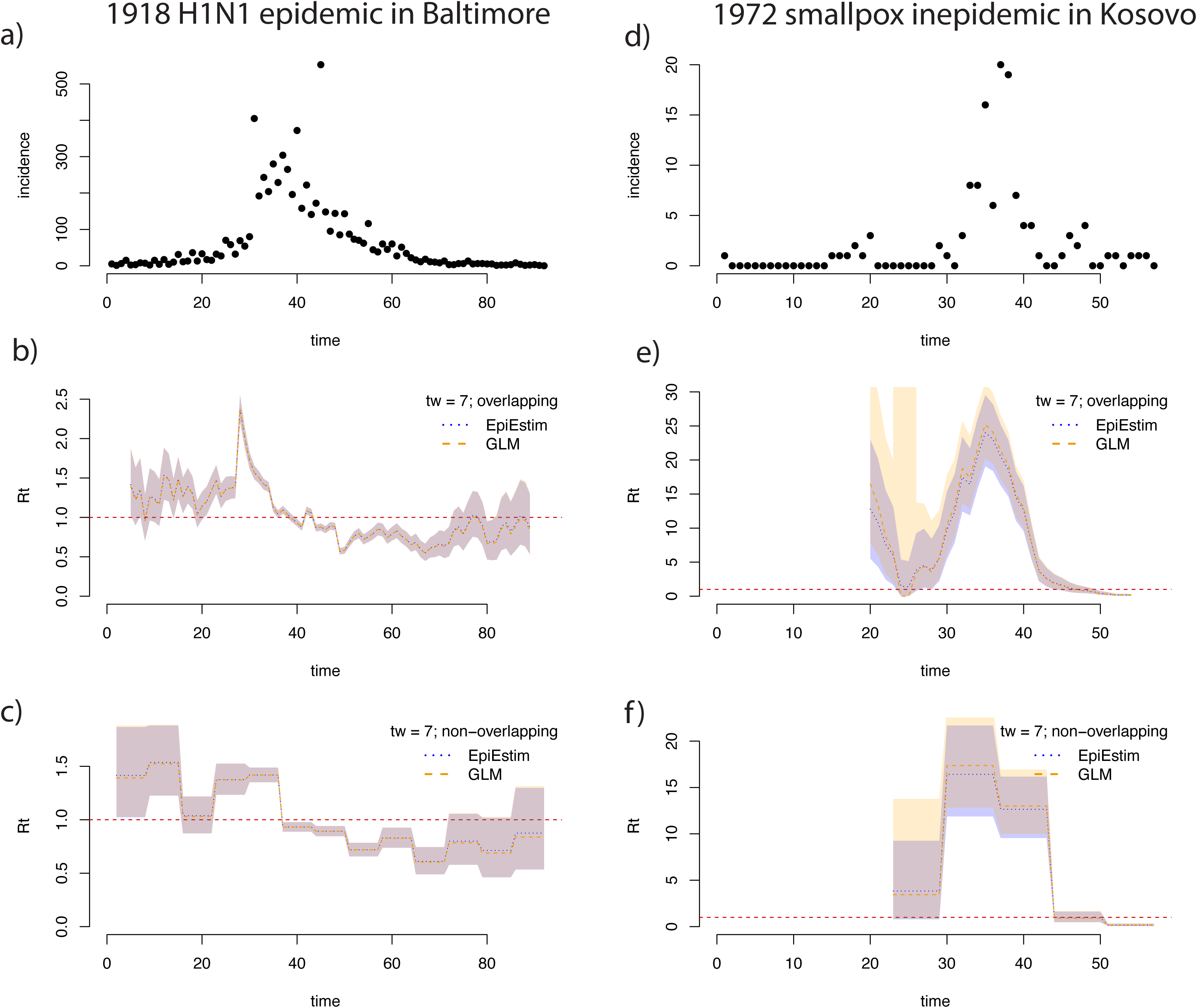
Reported incidence of influenza during the 1918 H1N1 epidemic in Baltimore (a), and the 1972 smallpox epidemic in Kosovo (d), dataset taken from the EpiEstim R-package. Estimates of *R_t_*’s over time using *EpiEstim* (blue) and *Rt.glm* (orange) assuming 7 days overlapping time-windows (b,e), plotted at the end of time-window. Blue and orange shading represent 95%Credible and Confidence Intervals respectively. Similar estimates are presented (c,f) assuming 7 days non-overlapping time-windows. Figures for other datasets and distinct time-window sizes are available in the SI.

Finally, using the 1918 influenza in Baltimore dataset (Figure SI.3), we demonstrated how using a smooth term to estimate *R_t_* resulted in broadly consistent trends with those presented here.

### b) Estimating temporal variation in Rt using GAM

Comparing the performances of the two frameworks in retrieving a simulated temporal trend, visually both *EpiEstim* and *Rt.glm* performed well (Fig.2, and Fig. SI.4-6). As expected, for *EpiEstim*, a reduced time-window led to wider credible intervals, while *Rt.glm* tended to yield to the narrowest confidence intervals. Overall, for both frameworks, poorer performance was associated with simulated incidences that included periods of very low incidence (Fig. SI.4-6). Relative to each other, low incidence affected *Rt.glm* estimates more than *EpiEstim* ones.

**Figure 2:**
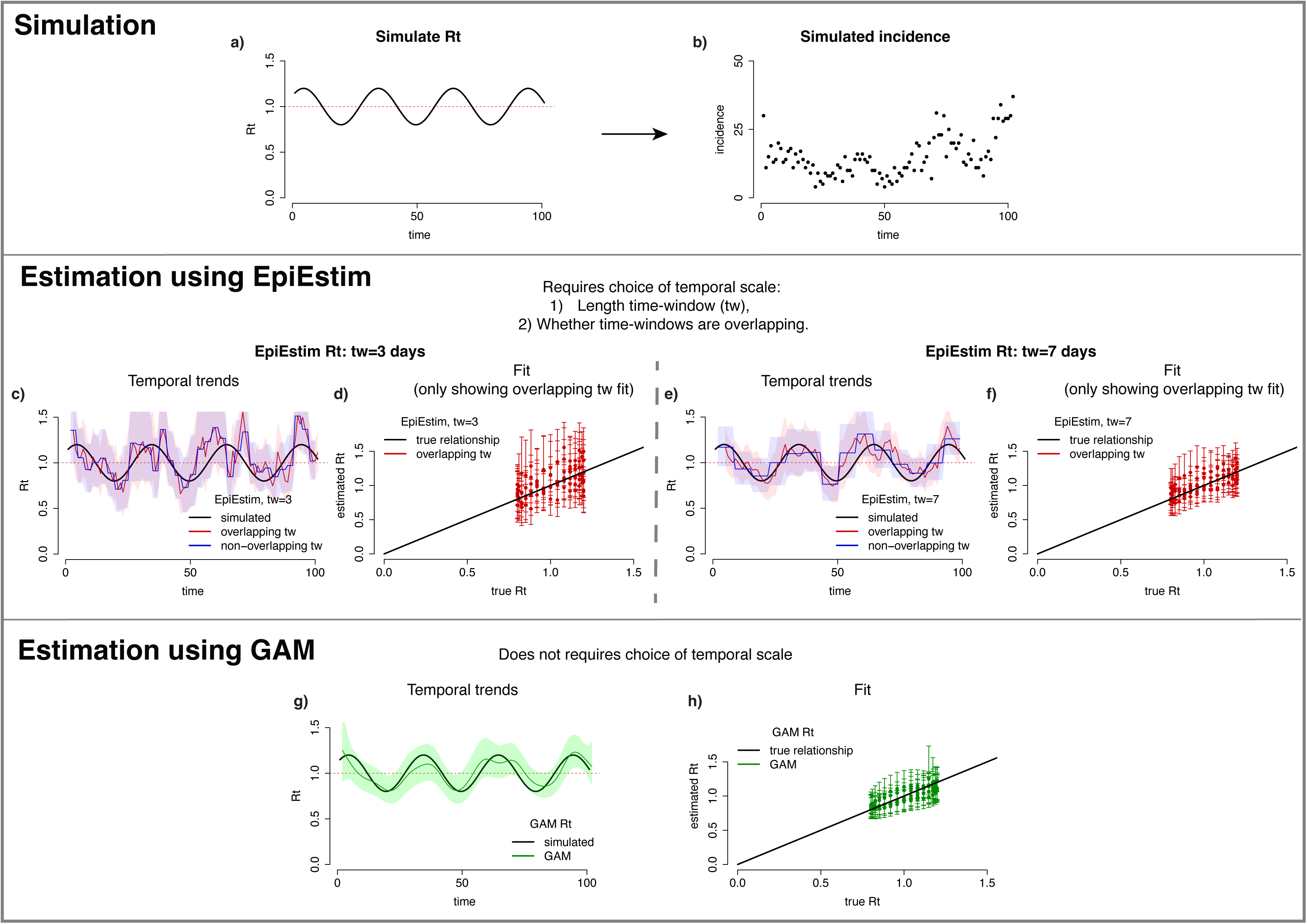
Simulated *R_t_* and incidence for one simulation (a,b). Using *EpiEstim* (c-f), we show estimates of *R_t_* over time (c,e) using overlapping (red) or non-overlapping (blue), as well as true *R_t_* (i.e. simulated, black). We also visualise performance by plotting estimated R_t_ against true *R_t_* (d,f). The estimated Rt assume either a 3 (c,d) or 7 days (e,f) time-window. During a time-window, *R_t_* is assumed constant. Using *Rt.glm* (g,h) and a GAM with a smooth temporal function, we also estimate *R_t_* over time (g), and show (h) the true *R_t_* (i.e. simulated) vs estimated *R_t_* (green). Note that *EpiEstim* users would need to use experience and judgment to choose the most appropriate time-window size.

Based on MSE and CRPS, median performance across simulations showed improved performance of *Rt.glm* over *EpiEstim* (Table 1, Fig. SI.7). The increase in performance was consistent across simulations (Table 2), such that *Rt.glm* performed better than any of *EpiEstim* estimates for well over 80% of simulations.

**Table 1:**
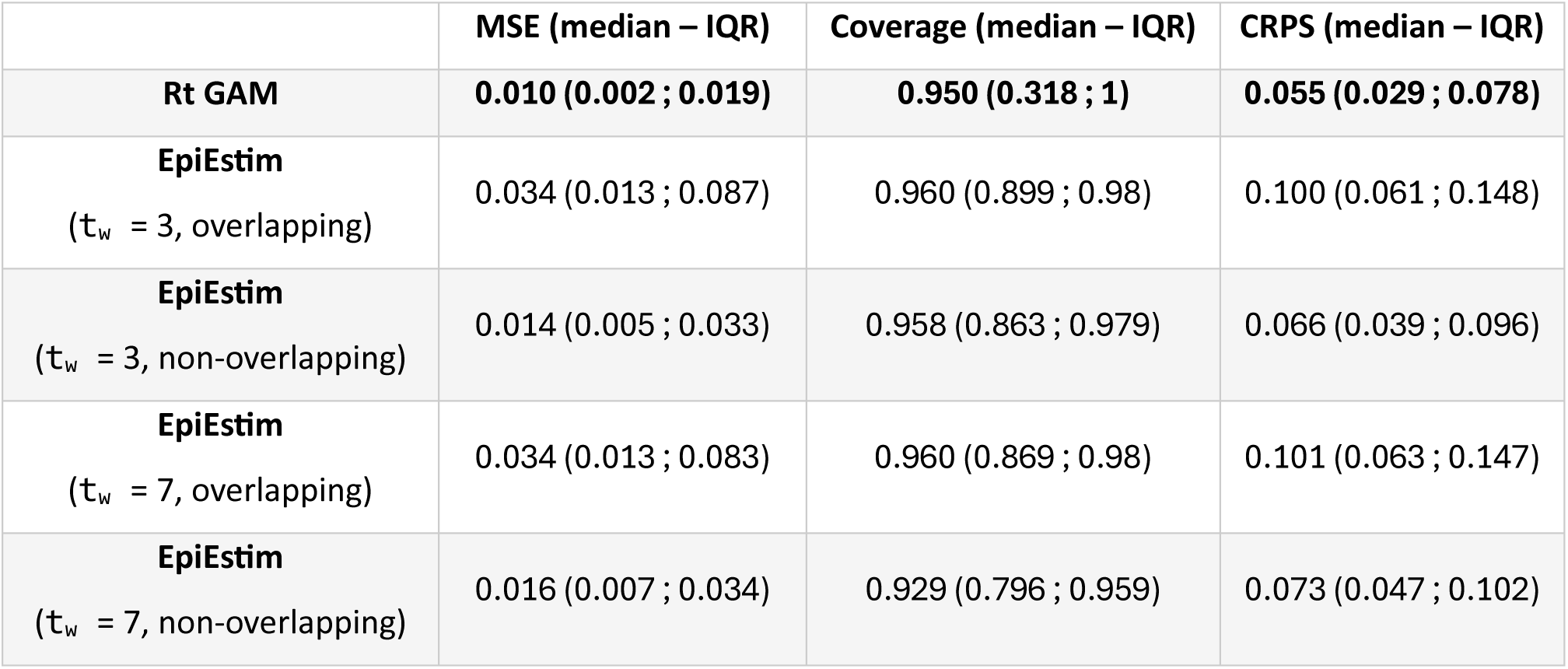
Performance of *EpiEstim* using distinct time-windows (3 vs. 7 days) and overlapping vs. non-overlapping against *Rt.glm*. Mean Square Error (MSE), % coverage of the 95% Credible/confidence interval (Coverage) and Continuous Ranked Probability Score (CRPS) were used as performance indicators. We show the median performance as well as the Interquartile Range (IQR) across 1000 simulations. Within a column (i.e. performance indicator), bold indicates the best performance.

**Table 2:** Percentage of simulations where *Rt.glm* performed better than any particular *EpiEstim* setup (i.e. distinct time-windows and overlapping characteristics), as well as across all EpiEstim estimates obtained. Performances were obtained for each performance indicator.

| Better performance of<br>Rt GAM vs. | % of MSE lower when<br>using GAM Rt | Coverage (% GAM Rt<br>closer to 95%) | % of CRPS lower<br>when using GAM Rt |
| --- | --- | --- | --- |
| **EpiEstim**<br>($t_w = 3$, overlapping) | 99.5 | 23.1 | 99.0 |
| **EpiEstim**<br>($t_w = 3$, non-overlapping) | 89.1 | 32.1 | 86.9 |
| **EpiEstim**<br>($t_w = 7$, overlapping) | 99.5 | 32.6 | 98.9 |
| **EpiEstim**<br>($t_w = 7$, non-overlapping) | 94.1 | 40.1 | 92.6 |
| **EpiEstim**<br>(across $t_w$, overlapping<br>settings) | 86.9 | 13.1 | 85.5 |

However, in some instances, *Rt.glm* tended to be marginally over-confident compared to *EpiEstim*, resulting in slightly lower coverage. While the median coverage across simulations performance was spot on, with 95.0% of estimated *R_t_*s including the true simulated *R_t_* within their 95%CI (i.e. % coverage, Table 1), in at least 60% of simulations (Table 2), *EpiEstim* outperformed *Rt.glm* in term of coverage (Table 2, Fig. SI.7). This was driven by simulations with low incidence resulting in *Rt.glm* failing to pick a temporal trend (Fig. SI.8-9) and essentially returning a single *R_t_* estimate throughout the simulation (or a very weakly temporally varying *R_t_*). Such estimates were characterised by narrow confidence intervals, driven by periods of higher incidence (Fig. SI.9). In contrast, for the same simulations, the ‘temporally agnostic’ *R_t_* estimates of *EpiEstim* returned the prior distribution of *R_t_* during periods of very low incidence, resulting in improved coverage (Fig. S.I9) (see SI4 for more details, including a brief discussion as to why switching the GAM error distribution to a negative binomial does not improve performance in those instances).

The results are shown here for a single simulated incidence. The simulated incidence was chosen as the one leading to the 25% best *Rt.glm* fit based on the ranked CRPS scores, similar figures based on median, 75% and worst *Rt.glm* fit are shown in SI (figs SI.4-6).

### c) Estimating Spatial variation in Rt using GAM

In retrieving a simulated spatial trend, while *Rt.glm* and *EpiEstim* with small spatial scale (UA spatial unit) performed well, using EpiEstim on aggregated incidence data (14 spatial units) led to poor performance (Fig. 3, and Fig. SI.11-13).

**Figure 3:**
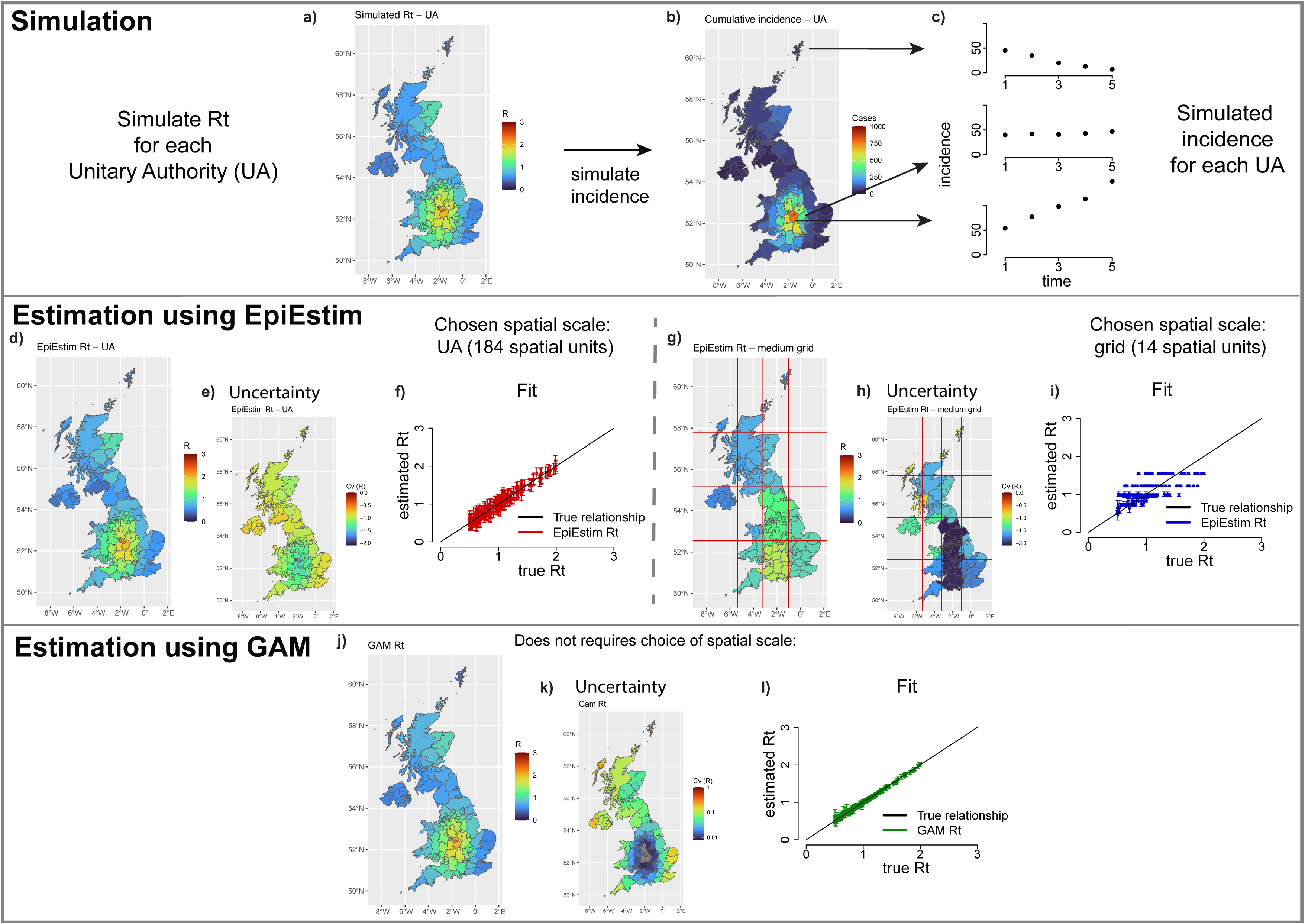
Simulated *R_t_*, cumulative incidence over the 5-days simulation, and selected daily incidence (a, b, c respectively). Maps of median estimated *R_t_* using *EpiEstim* (d, g). Estimated *R_t_* assume either independent *R_t_*s per UA (d, e, f) or homogeneity in *R_t_*s within a grid (red lines in g) of 14 regions (g, h, i). For each assumption scenario, we show maps of estimated *R_t_*s (d, g), as well as uncertainty, measured by the coefficient of variations (e, h), and true *R_t_*s (i.e. simulated) vs estimated *R_t_*s (f, i). Using *Rt.glm* (j, k, l), and a GAM formulation (i.e. with a smooth spatial function), we also estimate *R_t_*s over space (j), its uncertainties (k), and show true *R_t_*s (i.e. simulated) vs estimated *R_t_*s (j). Note that *EpiEstim* users would need to use experience and judgment to choose the most appropriate spatial scale.

Based on MSE and CRPS scores *Rt.glm* performed best overall (Table 3, Fig. SI.14), and this was true for all individual simulations (Table 4). The coverage score of *EpiEstim* at fine spatial scale (UA spatial unit) was marginally better than that of *Rt.glm* (median of 95.1% vs 93.4%, and see Fig. SI.13 for distributions). Again, *Rt.glm’*s worst scores, when using a spatial smoothing term, were associated with simulations that included very low incidence in multiple spatial units (Fig. SI.13).

**Table 3:** Similar to table 1, related to spatial estimation. Performance of *EpiEstim* using distinct spatial aggregation (UA vs. medium grid) against *Rt.glm*. Again, Mean Square Error (MSE), % coverage of the 95% Credible/confidence interval and Continuous Ranked Probability Score (CRPS) were used as performance indicators. We show the median performance as well as the Interquartile Range (IQR) across 10,000 simulations (i.e. 100 individuals kernel x 100 stochastic simulation per kernel). Within a column (i.e. performance indicator) bold indicates the best performance.

|  | MSE, median (IQR) | Coverage, median (IQR) | CRPS, median (IQR) |
| --- | --- | --- | --- |
| **Rt GAM** | **0.001 (0.001 ; 0.001)** | 0.934 (0.831 ; 0.962) | **0.017 (0.013 ; 0.019)** |
| **EpiEstim**<br>(UA grid) | 0.006 (0.005 ; 0.007) | **0.951 (0.918 ; 0.962)** | 0.045 (0.040 ; 0.046) |
| **EpiEstim**<br>(medium grid) | 0.065 (0.023 ; 0.079) | 0.251 (0.137 ; 0.295) | 0.174 (0.084 ; 0.198) |

**Table 4:** Similar to table 1, related to spatial estimation. Percentage of simulations where *Rt.glm* performed better than a particular *EpiEstim* setup (i.e. level of spatial aggregation), as well as across all EpiEstim estimates obtained. Performances were obtained for each performance indicator.

| Better performance of<br>Rt GAM vs. | % of MSE lower when<br>using GAM Rt | Coverage (% GAM Rt<br>closer to 95%) | % of CRPS lower<br>when using GAM Rt |
| --- | --- | --- | --- |
| **EpiEstim**<br>(UA grid) | 100 | 27.1 | 100 |
| **EpiEstim**<br>(medium grid) | 100 | 100 | 100 |
| **EpiEstim**<br>(across grid, i.e. the best of<br>either UA or medium grid) | 100 | 27.1 | 100 |

Therefore, as for temporal trends, 1) *Rt.glm* clearly outperformed *EpiEstim* in terms of MSE and CRPS performance, but 2) coverage can be marginally better for *EpiEstim* when incidence is low and a smoothing term is used.

The simulated incidence and fit shown led to the 25% best *Rt.glm* fit based on the ranked CRPS scores, similar figures based on median, 75% and worst *Rt.glm* fit are shown in SI.

## Discussion

We present a new method for estimating the time-varying reproduction number of infectious diseases (Rt). The method builds on commonly used branching process models and the renewal equation (7,10,16,17) and frames it within the framework of Generalized Linear Models (GLMs). Then, relying on an extension of GLMs, namely the Generalized Additive Models (GAMs)(23), we show how the novel method can address previous limitations in estimating the temporal and spatial granularity by allowing smooth estimation of *R_t_* variations over time and space.

The paper presents a proof-of-concept evaluation of our new framework and associated R-package, *Rt.glm*, by comparing its performance with that of the arguably most popular software tool for *R_t_* estimation, *EpiEstim* (16,21,30). The evaluation of performance relies on both historical datasets (e.g., influenza, SARS) and simulated outbreaks. Results demonstrate that *Rt.glm* provides promising improvements in estimating *R_t_* by effectively capturing temporal and spatial heterogeneities in transmission dynamics. The improvement is marked in terms of accuracy, based on the lower Mean Square Error. While the characterisation of uncertainty was reasonable (based on coverage of confidence intervals), when using a smooth, *Rt.glm* showed slight overconfidence in its estimates when incidence was very low. Overall, on the balance of accuracy and capturing uncertainty (as measured by the Continuous Ranked Probability Score)(26,27), *Rt.glm* clearly outperformed EpiEstim in every setting explored.

We further stress that the issue of marginal overconfidence was only linked to situation where i) a smooth was used, and ii) incidence was low. In such situation, we demonstrate that the smooth *R_t_* estimates lacked temporal or spatial variability. While one might suggest using an over-dispersed distribution in the inference (e.g. negative binomial regression instead of Poisson), we demonstrate that this would only compound the problem, by effectively reducing an already weak temporal/spatial signal. In such situation, we recommend that no smooth is used, instead relying on independent time-window to infer transmissibility (i.e. *Rt.glm* parameterisation equivalent to *EpiEstim*). Nevertheless, i) the implementation of an over-dispersed distribution is supported within *Rt.glm*, and ii) can be used with or without a smoothing term, i.e. resulting in a generalisation of *EpiEstim* with an underlying over-dispersed distribution.

The proposed *Rt.glm* R-package simplifies the implementation of *R_t_* estimation by offering a user-friendly tool for researchers and public health professionals. In particular, it largely removes the need for users to subjectively choose the temporal and/or spatial scales at which *R_t_* is assumed to be homogeneous (i.e. when using a smoothing term, but see section SI1). Additionally, using *Rt.glm*, and relying on GAM for fitting, users can estimate the effective degree of freedom (a measure of complexity) associated with the smooth term(s), which inform on the characteristic time / spatial window of *R_t_* fluctuations. This framework holds promise as a robust alternative for real-time *R_t_* estimation and forecasting, advancing epidemic monitoring and response efforts. The framework can be viewed as an extension of *EpiEstim*, as its results reduce to those of *EpiEstim* in its simplest application, e.g. without using smoothed parameters. The framework can also reproduce some, but not all, of *EpiEstim* features. For example, as *EpiEstim*, it can estimate *R_t_* under the assumption of overlapping time-windows(16), but unlike *EpiEstim*, *Rt.Glm* currently is unable to account for uncertainty in the SI distribution (*EpiEstim* can, relying on contact tracing data for instance)(21), nor reconstruct daily incidence from temporally aggregated data (31), or estimate the transmission advantage a variant (32). However, the novel framework opens new avenues for future methodological improvements and real-world applications.

First, by relying on GAMs, it provides a robust foundation for integrating spatio-temporal variations. This can enhance precision in estimating transmission dynamics across diverse geographic areas and temporal scales. This would be especially important during heterogeneous outbreaks (33–35).

Second, GLMs have proven extremely useful in characterising the links between multiple covariates and dependent variables (36,37). In an infectious disease context, this paves the way to assess drivers of transmission by constraining *Rt* estimates to covariates, i.e. putative drivers of transmission dynamics. Critically, this includes evaluating the impact of control interventions on transmissibility, an area of research that is not only critical to guide public health response (20) but also challenging to perform, both theoretically and computationally, with currently existing methods (38–41). Furthermore, the ability to incorporate smooth, non-linear, functions for temporal and spatial predictors would support a more nuanced understanding of complex epidemic patterns.

Third, future extensions could also focus on improving the characterization of uncertainty and overdispersion in *Rt* estimates, i.e. by using a negative binomial distribution instead of the default Poisson distribution (as already mentioned above). Importantly, while the implications and fundamental rationale for using overdispersion are non-trivial (see below), practical implementation with *Rt.glm* is itself trivial, as supported within GLM/GAM frameworks. Furthermore, it could be implemented with or without smooth, leading to a generalisation of *EpiEstim* with an underlying over-dispersed distribution. Incorporating these aspects would make the framework more robust for data with high variability (e.g. variability in transmission or reporting), where traditional methods, often relying on an assumption of Poisson offspring distribution, often struggle. We briefly explore this in SI (see SI section 4), and highlight the non-trivial nature of the problem, e.g. while using a negative binomial as a default might seem a reasonable approach, we would question and caution how and when to use it, especially when using a smoothing term in situation of low incidence (see SI4 for further discussion and recommendations). Nevertheless, since the early 70s, General Linear Models were formally extended to Generalized Linear models to allow more flexibility in the underlying assumed distribution of the dependent variable (37), including developments to allow more flexibility in error structure. Substantial statistical literature and tools have been developed to handle heterogeneities robustly and accurately, including heterogeneities in residuals (36,37). In the context of infectious diseases, this paves the way to test for and estimate overdispersion in incidence, and perhaps mechanistically link, and estimate, characteristics of the underlying generative processes such as superspreading (33). Future work will be required to carefully introduce the use of over-dispersed distribution (e.g. negative binomial, quasi-Poisson, Tweedie), and such work will need to either tackle this phenomenologically (e.g. using what is often referred to as either the NB1 or NB2 models (42)), or mechanistically (e.g. after formally disentangling the consequences of various sources of heterogeneities in the expected variance of observed incidence.

Fourth, the extension of the GLMs to account for random effects linked to either experimental or observational data collection design (i.e. mixed effect models) has proven increasingly popular to refine estimates and better account for the partitioning of variance associated with particular groups (43). This means the field of *R_t_* estimation could join the fields of ecology, non-communicable epidemiology and many others in their ubiquitous use and application of mixed effect models.

A full listing of further possible developments is outside the scope of this proof-of-concept paper, and will require further validation and package development, but there are certainly many more than the anticipated extensions discussed above. GLMs have a long and rich history, which also involves practical issues around model selection, comparison and averaging, as well as theoretical issues around causality (i.e. causal inference) that could be more readily explored with our novel framework relying on past and current developments of GLMs.

In conclusion, the *Rt.GLM* framework presents a substantial and promising advancement in the estimation of time-varying reproduction numbers, addressing key limitations of existing methodologies by integrating spatio-temporal flexibility within a statistically robust framework. By enabling smoother and more accurate estimations of Rt, the approach has the potential to improve the understanding of transmission dynamics and the evaluation of interventions across diverse settings. While challenges remain, such as improving uncertainty characterization at low incidence levels when using a smoothing term, the framework provides a strong foundation for future developments, including the incorporation of covariates and mixed effects. Overall, *Rt.GLM* offers a versatile and user-friendly tool to enhance epidemic monitoring and public health decision-making.

## Data Availability

All data produced in the present study are available upon reasonable request to the authors

## Acknowledgements

I would like to thank Anne Cori, Christl Donnelly and Johannes Bracher for helpful discussions and comments to improve this manuscript.

## Author contributions: CRediT

Sole author contribution to conceptualisation, formal analysis and writing.

